# Effects of Government Mandated Social Distancing Measures on Cumulative Incidence of COVID-19 in the United States and its Most Populated Cities

**DOI:** 10.1101/2020.05.22.20110460

**Authors:** Bhuma Krishnamachari, Alexander Morris, Diane Zastrow, Andrew Dsida, Brian Harper, Anthony J. Santella

## Abstract

COVID-19, caused by the SARS-CoV-2 virus, has quickly spread throughout the world, necessitating assessment of the most effective containment methods. Very little research exists on the effects of social distancing measures on this pandemic. The purpose of this study was to examine the effects of government implemented social distancing measures on the cumulative incidence rates of COVID-19 in the United States on a state level, and in the 25 most populated cities, while adjusting for socio-demographic risk factors. The social distancing variables assessed in this study were: days to closing of non-essential business; days to stay home orders; days to restrictions on gathering, days to restaurant closings and days to school closing. Using negative binomial regression, adjusted rate ratios and 95% confidence intervals were calculated comparing two levels of a binary variable: “above median value,” and “median value and below” for days to implementing a social distancing measure. For city level data, the effects of these social distancing variables were also assessed in high (above median value) vs low (median value and below) population density cities. For the state level analysis, days to school closing was associated with cumulative incidence, with an adjusted rate ratio of 1.59 (95% CI:1.03,2.44), p=0.04 at 35 days. Some results were counterintuitive, including inverse associations between cumulative incidence and days to closure of non-essential business and restrictions on gatherings. This finding is likely due to reverse causality, where locations with slower growth rates initially chose not to implement measures, and later implemented measures when they absolutely needed to respond to increasing rates of infection. Effects of social distancing measures seemed to vary by population density in cities. Our results suggest that the effect of social distancing measures may differ between states and cities and between locations with different population densities. States and cities need individual approaches to containment of an epidemic, with an awareness of their own structure in terms of crowding and socio-economic variables. In an effort to reduce infection rates, cities may want to implement social distancing in advance of state mandates.

## Introduction

The novel coronavirus, known as SARS-CoV-2, was recently identified as the causative agent of COVID-19 (1). The virus, originating from Wuhan, China, quickly spread throughout the world, leaving countries scrambling to assess the most effective containment methods (2). Though the virus initially presented in the United States (U.S.) after infected individuals came into the country from international locations, the mode of transmission quickly became person to person (3). According to one source, some states, such as Washington, California, and New York were affected at high rates (4). It should be noted that since this article was published, the states with the highest rates have changed (5). Similarly, some cities were disproportionately affected, with New York City (NYC), emerging as a “hotspot” when incidence rates climbed higher and faster than other locations (6).

As the world started to learn more about the biology of SARS-CoV-2, proposals for containment measures attempted to address the modes of transmission. Spread of SARS-CoV-2 occurs primarily through respiratory droplets (7). However, the virus can also survive on hard surfaces for days (8). Because the spread of the virus is largely contingent upon proximity to one another, common containment measures included attempts to create physical distance (e.g. public health measures such as restrictions on gatherings) (9). As the U.S. scrambled to establish containment measures, states and local governments laid out vastly heterogeneous rules on how individuals would prevent close contact (10,11).

There is little research on the effect of social distancing on the pandemic. A report from the Centers for Disease Control and Prevention examined data from the metropolitan areas of San Francisco, Seattle, New Orleans and NYC in terms of the timing of policy measures, community mobility, and incidence rates. They found that the percentage of residents leaving home declined as the number of policies issued increased, with data trends suggested that social distancing measures may decrease incidence rates. The analysis was limited by lack of information on confounders (e.g. age, gender, and race) and limited data on mobile phone coverage (12). Another recent publication looked at the impact of several social distancing measures on the growth rate of confirmed COVID-19 cases using county level data across the U.S. They reported that government-imposed social distancing measures reduced the daily growth rate by 5.4 % after 1–5 days, 6.8 % after 6–10 days, 8.2% after 11–15 days, and 9.1% after 16–20 days, and suggested that failure to enact these types of restrictions would fuel exponential spread (13). Though there are not many published studies on the effects of social distancing on COVID-19, social distancing measures associated with other epidemics for Severe Acute Respiratory Syndrome (SARS) outbreak and Ebola virus disease have been reported, but comparisons are difficult because those viruses frequently affected a different demographic (children) than COVID-19 (14,15).

Most current published studies utilize mathematical modeling to make predictions based on hypothetical situations, yielding a variety of conclusions. One study modeled the effects of social distancing measures on the progression of the COVID-19 epidemic in Wuhan, China, which showed that physical distancing measures may be most effective if delayed rather than done as a sudden lifting of interventions. However, they noted that results varied by the duration of infectiousness and the role of school children in the epidemic (16). Another group created a model to study the impact of social distancing and school closure on viral transmission while accounting for age differences. They reported that social distancing in China during their outbreak was sufficient to control COVID-19, with a 40-60% reduction in peak incidence with proactive school closures (17). A recent mathematical model used to investigate the effectiveness of social distancing interventions in a mid-sized U.S. city (modeled after Seattle) showed that interventions that start earlier in the epidemic delay the epidemic curve while interventions initiated later flatten the epidemic curve (18).

The purpose of the present study was to examine the effects of social distancing measures on cumulative incidence rates of COVID-19, while taking numerous other risk factors into consideration. Specifically, using real world evidence sources, we investigated the differences between the states as well as the 25 most populated cities in the U.S. in the effectiveness of government mandated social distancing measures.

## Methods

Data was gathered for the 50 states and the District of Columbia and for the 25 largest cities in the United States. The District of Columbia was analyzed both as a state and a city. Data for cumulative counts of cases was obtained from the Johns Hopkins University COVID-19 Dashboard(5). Data for dates of social gathering restrictions for each state was acquired from the University of Washington GitHub dataset (19). The social distancing variables gathered were days to closing of non-essential business; days to stay home orders; days to restrictions on gathering, days to restaurant closings and days to school closing. Data at the city level was collected from each city’s government website. All socio-demographic variables were acquired through the Census (20, 21). Data regarding the prevalence of health conditions was obtained from the Centers for Medicare and Medicaid (22).

### Statistical Analyses

All analyses were done for states and cities separately. Cumulative incidence rates were calculated as the number of reported cases at a given time divided by the population size of that location, multiplied by 100,000. The first day of the epidemic at any location was defined as the first day at least 50 cases were present. The days to implementing each social distancing measure were calculated as the dates between emergency declaration to implementation of the measure. The days of preparation time available for each location were calculated as the number of days from the first case reported in the Johns Hopkins University data to the first day at least 50 cases were present in that location.

Prevalence of several variables were evaluated for confounding, including percentage in each location of people who are: Black/African American; Hispanic; age 65 and up; female; in poverty; have a college degree; own a computer; have internet access at home; use carpooling for work; use public transit for work; or have one of the following medical conditions: asthma; diabetes; chronic obstructive pulmonary disease (COPD); hypertension; heart failure or ischemic heart disease. A binary variable created for medical conditions using a cutoff of being above median or median and below values for all medical conditions combined. For analysis of state data, percentage of the total population represented by urban population and population density were evaluated using data from the Census (20, 21).

The median, mean and standard deviation (SD) were reported for the days to implementation of each social distancing variable and other variables that may be social distancing predictors. These variables were converted into binary variables for analysis, using the median value in the study population as a cutoff. Variables were created separately for states and for cities using their respective median values.

Using negative binomial regression, rate ratios and 95% confidence intervals were calculated comparing the “above median value,” and “median value and below” groups at days 14, 21, 28, 35 and 42 of the epidemic. Day 7 was not included in the analysis because reporting of cases was initially erratic. Goodness-of-fit for including different confounders was assessed using Akaike’s Information Criterion (AIC)-based selection criteria to create a final model of predictors. A sensitivity analysis was conducted by re-running the analysis with omission of California, Illinois, New York, Pennsylvania and Texas as states that have the five most populous cities. An additional sensitivity analysis was run excluding California, District of Columbia, Massachusetts, New York and Washington state since they hold the top five cities with highest population densities. Finally, a sensitivity analysis was run on city data by analyzing with omission of each city.

A sub-analysis was conducted for the city data to examine the effects of social distancing measures in cities of high population density (above median value) vs lower population density (median and below). A four-level categorical variable was created using the binary population density variable and each binary social distancing variable. Day 35 cumulative incidence was used for analysis because it represents a time point far enough into the epidemic to detect a pattern, but not so recent that data may still be getting updated (for e.g. 42 week data). Cumulative incidence was log transformed in order to normalize data for analysis. All comparisons were made between four groups: Low-Low: Low population density and median and below value for variable being assessed, Low-High: low population density and above median value for variable being assessed, High-Low: high population density and median or below value for variable being assessed and High-High: high population density and above median value for variable being assessed. Both ANOVA with Bonferroni correction and the Kruskal Wallis test were used to evaluate the relationship between the four-level categorical variables and log cumulative incidence. Box plots were constructed to view the median cumulative incidence values for the different social distancing categories that exhibited significant results on both the ANOVA and Kruskal Wallis tests. Statistical analysis was conducted using SAS 9.4. Statistical significance was set at p-value ≤0.05.

## Results

The mean and median values for government implemented factors and possible confounders are shown in Table 1. Table 2 shows the differences between states that took above median vs median and below number of days to implementing different social distancing measures. Adjustment for days for preparation, population density percentage of total population represented by urban population, percent female, 65 and over and percent Black/African American resulted in the best model fit. Days to closure of non-essential businesses was inversely associated with cumulative incidence, with rate ratios consistently between 0.49 to 0.66. Days to restrictions on gatherings was also inversely associated with cumulative incidence, with rate ratios between 0.48 to 0.64, but after adjustment, none of these associations remained statistically significant. Days to school closing was associated with cumulative incidence on day 35 and 42, with an adjusted rate ratio of 1.59 (95% CI:1.03,2.44), p=0.04 at 35 days, and adjusted rate ratio of 1.64 (95% CI:1.07,2.52), p=0.04 at 42 days. Sensitivity analysis with omission of states that may skew results did not alter results.

**Table 1:** Social Distancing and Confounding Variables, Mean and Median % Values

|  | States<br>N=50 |  | Cities<br>N=25 |  |
| --- | --- | --- | --- | --- |
|  | Mean (SD) | Median | Mean (SD) | Median |
| **Social Distancing Factors** |  |  |  |  |
| Days to closing of non-essential business | 26 (15) | 19 | 28 (14) | 20 |
| Days to Stay home orders | 19 (10) | 17 | 14 (5) | 14 |
| Days to Restrictions on gathering | 10 (11) | 8 | 13 (14) | 8 |
| Days to Restaurant closings | 9 ( 8) | 8 | 17 (17) | 9 |
| Days to School closing | 7(7) | 6 | 8 (9) | 7 |
| Population Density per Square Mile | 385 (1377) | 101 | 6361 (5965) | 3922 |
| Percent who use Public Transit to get to work | 4 (6) | 1.4 | 3 (1) | 2.20 |
| Percent who use Carpooling to get to work | 9 (1) | 9 | 6 (7) | 2 |
| **Demographic Factors** |  |  |  |  |
| 65 and older | 16 (2) | 16 | 12 (1) | 12 |
| Female | 51% (1%) | 51 | 51% (1%) | 51 |
| Black/African American | 11% (11%) | 8 | 21 (18) | 23 |
| Hispanic | 12 (10) | 10 | 27 (19) | 29 |
| Poverty | 12 (3) | 13 | 18 (5) | 19 |
| **Medical Conditions** |  |  |  |  |
| Diabetes | 26 (3) | 26 | 28 (5) | 29 |
| Asthma | 5 (1) | 5 | 6 (1) | 6 |
| Heart Failure | 13 (2) | 13 | 14 (3) | 14 |
| Hypertension | 55 (7) | 57 | 55 (7) | 57 |
| Ischemic Heart Disease | 25 (4) | 25 | 25 (5) | 25 |
| COPD | 11 (2) | 11 | 10 (2) | 9 |
| Above median value for all diseases (categorical) | n=21 (41%) |  | n=12 (48%) |  |

**Table 2:** Effect of Social Distancing on States

|  | 14 days<br>Rate SD<br>Rate Ratio (95% CI) | 21 days<br>Rate SD<br>Rate Ratio (95% CI) | 28 days<br>Rate SD<br>Rate Ratio (95% CI) | 35 days | 42 days |
| --- | --- | --- | --- | --- | --- |
| All states avg rate | 32.57 (20.78) | 77.38 (63.59) | 135.90 (129.75) | 196.42 (195.48) | 260.02 (259.57) |
| **Closing of NE Business** |  |  |  |  |  |
| Avg rate median or below n=26 | 40.49 (24.30) | 102 (76.64) | 181.50 (159.70) | 196.42 (195.48) | 341.10 (320.00) |
| Avg rate above median n=25 | 24.34 (12.08) | 51.63 (30.82) | 88.48 (62.56) | 261.10 (241.60) | 175.70 (138.50) |
| Unadjusted rate ratio | **0.60 (0.44, 0.81) p=0.001** | **0.51 (0.36,0.70) p< 0.0001** | **0.49 ( 0.34, 0.71) p=.0002** | **0.49 (0.33,0.73) p=0.0004** | **0.51 (9 0.34, 0.77) p=.0013** |
| Adjusted Rate ratio rate* | **0.60 ( 0.46, 0.78) p=.0002** | **0.56(0.40,0.78) p=.0007** | **0.60 (0.42, 0.87) p=0.0065** | **0.66 (0.45, 0.96) p=.0297** | 0.71 (0.48, 1.05) p=.08 |
| **Stay at Home Orders** |  |  |  |  |  |
| Avg rate median or below n=27 | 33.37 (20.14) | 83.44 (69.92) | 148.60 (151.80) | 129.20 (97.77) | 275.80 (304.60) |
| Avg rate above median n=24 | 31.67 (21.88) | 70.57 (56.32) | 121.60 (100.80) | 212.20 (229.80) | 242.30 (202.40) |
| Unadjusted rate ratio | 0.95 (0.68, 1.32) p=.75 | 0.85 ( 0.58, 1.23) p=.38 | 0.82 ( 0.54, 1.23) p=.34 | 0.84 ( 0.55, 1.29) p=.43 | 0.90 ( 0.60, 1.35) p=.60 |
| Adjusted Rate ratio rate* | 0.75 (0.55,1.03) p=.07 | 0.75 (0.52,1.09) p=.12 | 0.81 ( 0.55,1.22) p=.32 | 0.90 ( 0.60, 1.35) p=0.60 | 1.00 (0.64 , 1.47) p=.88 |
| **Restrictions on Gatherings** |  |  |  |  |  |
| Avg rate median or below n=30 | 35.71 (23.66) | 90.74 (76.87) | 166.20 (158.10) | 178.60 (150.80) | 330.10 (313.10) |
| Avg rate above median n=21 | 27.29 (13.72) | 58.30 (29.85) | 92.65 (50.02) | 245.50 (237.60) | 160.0 (91.93) |
| Unadjusted rate ratio | 0.82 (0.59,1.142) p=.24 | **0.64 ( 0.44,0.93) p=.02** | **0.56 (0.38,0.83) p=.004** | **0.51 ( 0.34,0.77) p=.001** | **0.48 ( 0.32,0.73) p=.0005** |
| Adjusted Rate ratio rate* | 0.85 (0.61,1.18) p=.34 | 0.78 (0.52,1.16), p=.23 | 0.75 ( 0.50, 1.134) p=.17 | 0.73 ( 0.48, 1.10) p=.13 | 0.69 (0.46, 1.04) p=.07 |
| **Closing Restaurants** |  |  |  |  |  |
| Avg rate median or below n=32 | 35.71 (23.66) | 85.50 (71.25) | 150.10 (140.20) | 215.60 (201.70) | 285.80 (265.80) |
| Avg rate above median n=19 | 27.29 (13.72) | 63.72 (46.62) | 111.90 (109.20) | 164.00 (185.30) | 216.60 (249.70) |
| Unadjusted rate ratio | 0.77 (0.55,1.07) p=.12 | 0.75 ( 0.51, 1.09) p=.13 | 0.75 (0.49, 1.14) p=.17 | 0.76 (0.49, 1.18) p=.22 | 0.76 (0.48, 1.19) p=.22 |
| Adjusted Rate ratio rate* | 0.95 ( 0.69, 1.30) p=.73 | 0.80 ( 0.55, 1.16) p=.24 | 0.72 (0.49,1.07) p=.10 | 0.70 (0.47, 1.03) p=.07 | 0.67 (0.45,1.00) p=.05 |
| **Closing Schools** |  |  |  |  |  |
| Avg rate median or below n=26 | 34.36 (22.24) | 71.68 (60.51) | 113.90 (103.10) | 153.50 (136.40) | 194.90 (171.20) |
| Avg rate above median n=25 | 30.71 (19.43) | 83.31 (67.36) | 158.80 (151.40) | 241.10 (47.39) | 327.70 (317.00) |
| Unadjusted rate ratio | 0.89 (0.64,1.24) p=.51 | 1.16 ( 0.80,1.69) p=.43 | 1.40 ( 0.93,2.09) p=.11 | **1.57 ( 1.04, 2.37) p=.03** | **1.68 (1.11, 2.55) p=.01** |
| Adjusted Rate ratio rate* | 1.17 ( 0.80, 1.70) p=.42 | 1.38 (0.91, 2.10) p=.13 | 1.52 (0.98, 2.33) p=.06 | **1.59 (1.03,2.44) p=.04** | **1.64 (1 .07, 2.52) p=.02** |

\*Adjustment for days for preparation, population density, Percentage of total population represented by urban population, percent Female, 65 and over, percent black

Table 3 shows the differences between cities that took above median vs median and below number of days to implementing different social distancing measures. The best model fit included adjustment for percent of Black/African American individuals in a city. Days to closure of non-essential businesses was inversely associated with cumulative incidence at days 28, and 42, with adjusted rate ratios of 0.64 and 0.63, respectively. Increased days to restrictions on gatherings was statistically significant (rate ratio=2.17 (95% CI: 1.14,4.12) p=0.02), but after adjustment for the percent black individuals in a city, the result was no longer statistically significant (rate ratio=0.97 (0.60,1.56) p=0.90). Population density was consistently associated with increased cumulative risk at all time points, with an adjusted rate ratio of 2.35 (95% CI: 1.50, 3.65) p=0.0002) at 42 days.

**Table 3:**
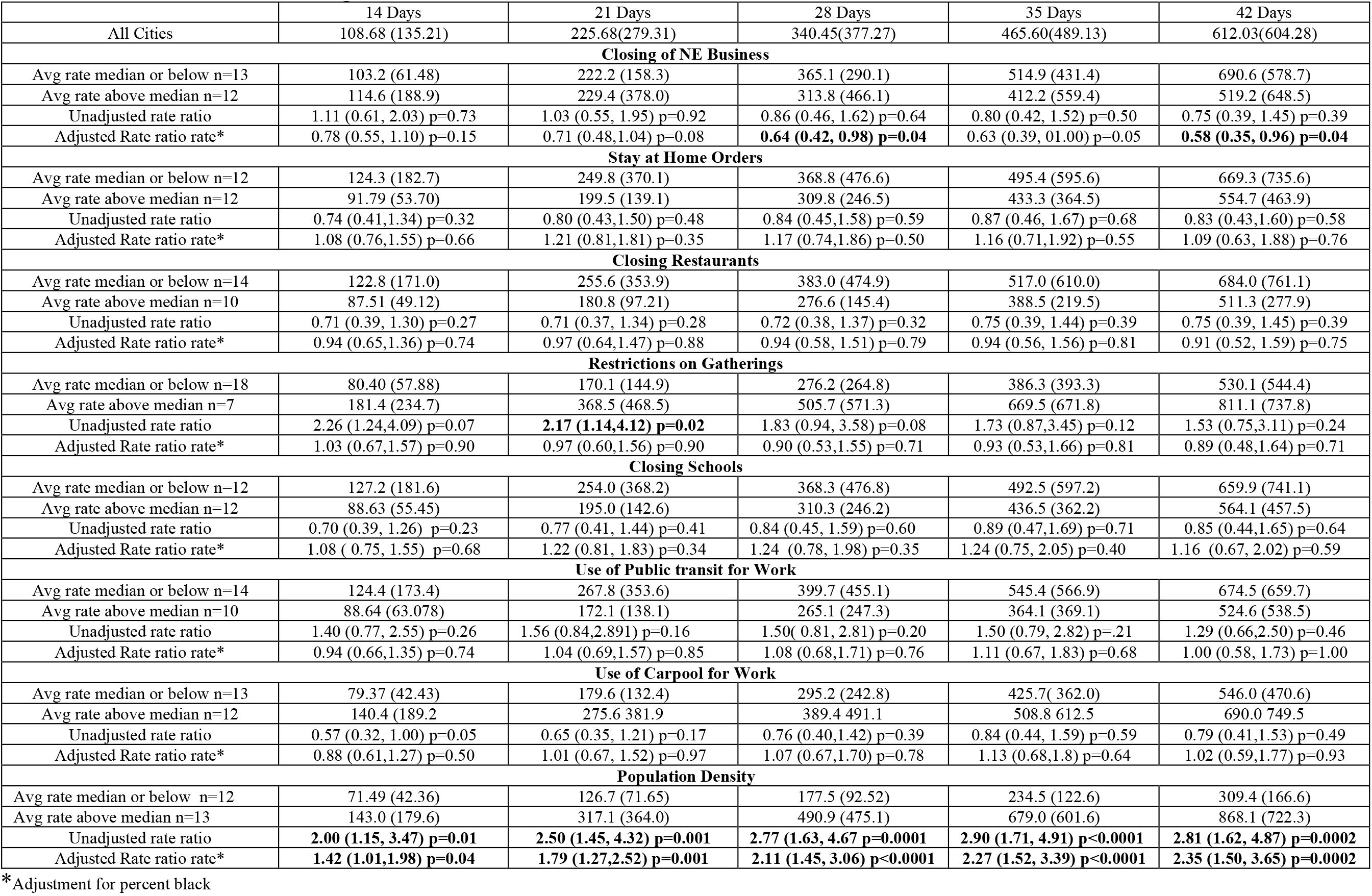
Effect of Social Distancing on Cities

Table 4 displays the statistically significant results for both ANOVA and Kruskal-Wallis tests when evaluating cumulative incidence at day 35, looking at the combined effects of various factors and population density. For days to restaurant closure, there was a statistically significant difference in the cumulative incidence between the Low-High group (median or below days to implementation in high population density cities) and the Low-Low group (median or below days to implementation in low population density cities). For days to school closure there was a statistically significant difference in the cumulative incidence between the Low-High group (median or below days to implementation in high population density cities) and the Low-Low group (median or below days to implementation in low population density cities). For restrictions on gatherings, there was a statistically significant difference in the cumulative incidence between the High-High group (above median to implementation in high population density cities) and the Low-Low group (median or below days to implementation in low population density cities). Figure 1 shows the box plots for the three social distancing measures/population density combinations that had statistically significant results using both ANOVA and the Kruskal-Wallis tests.

**Figure 1.**
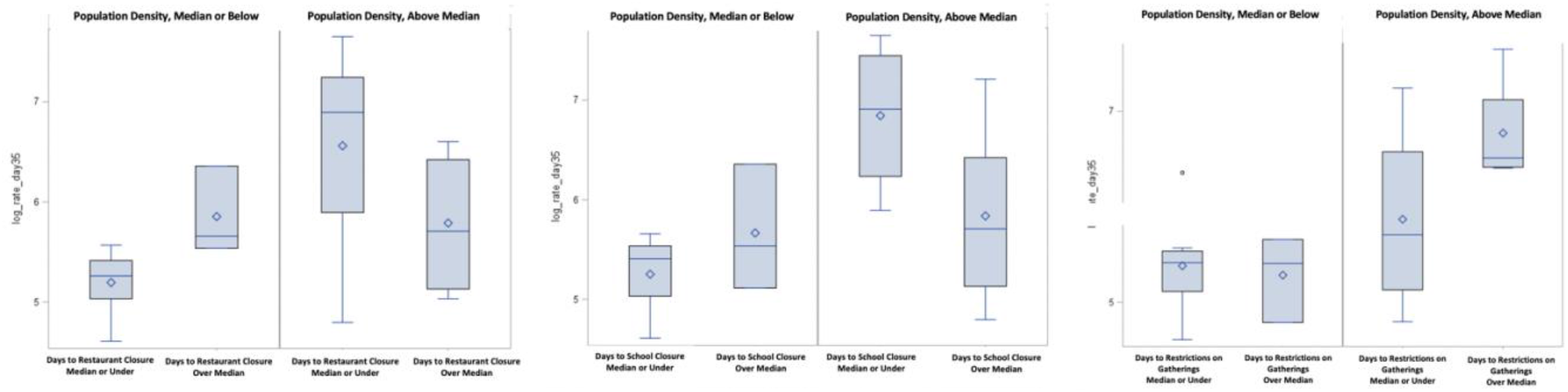
Social Distancing Measures and Population Density: Boxplots of Log Transformed Cumulative Incidence

**Table 4:** Variable Effects on Cumulative Incidence in Low and High Density Cities* at Day 35

| Comparisons were made between 4 groups:<br>-Low-Low: Median or below value for social distancing variable and low population density<br>-High-Low: Above median value for social distancing variable and low population density<br>-Low-High: Median or below value for social distancing variable and high population density<br>-High-High: Above median value for social distancing variable and high population density |  |  |  |
| --- | --- | --- | --- |
|  | ANOVA p-value | Kruskal-Wallis p-value | ANOVA significant differences |
| Days to Restaurant Closure | 0.006 | 0.034 | Low-High as compared to Low-Low |
| Days to School Closure | 0.006 | 0.034 | Low-High as compared to Low-Low |
| Days to Restrictions on Gatherings | 0.036 | 0.048 | High-High as compared to Low-Low |

\***Low population density:** Houston, TX; Phoenix, AZ; San Antonio, TX; Dallas, TX; Austin, TX; Jacksonville, FL; Fort Worth, TX; Columbus, OH; Charlotte, NC; Indianapolis, IN; El Paso, TX; Nashville, TN. **High population density:** New York City, NY; Los Angeles, CA; Chicago, IL; Philadelphia, PA; San Diego, CA; San Jose, CA; San Francisco, CA; Seattle, WA; Denver, CO; Washington, D.C.; Boston, MA; Detroit, MI; Portland, OR.

Table 5 shows the cumulative incidence rates for the social distancing measures and population density combinations that had statistically significant results using both ANOVA and the Kruskal-Wallis tests.

**Table 5:**
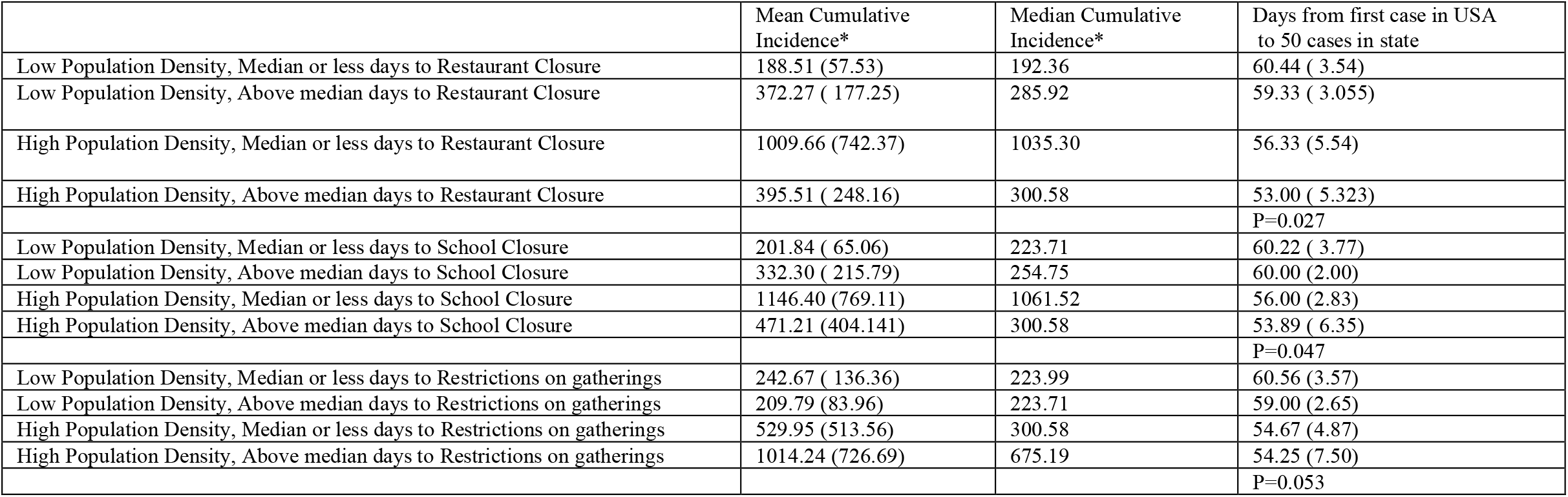
Day 35 Cumulative Incidence Rates in Low and High Density Cities, Categorized by Days to Social Distancing Measures

|  | Mean Cumulative Incidence* | Median Cumulative Incidence* | Days from first case in USA to 50 cases in state |
| --- | --- | --- | --- |
| Low Population Density, Median or less days to Restaurant Closure | 188.51 (57.53) | 192.36 | 60.44 ( 3.54) |
| Low Population Density, Above median days to Restaurant Closure | 372.27 ( 177.25) | 285.92 | 59.33 ( 3.055) |
| High Population Density, Median or less days to Restaurant Closure | 1009.66 (742.37) | 1035.30 | 56.33 (5.54) |
| High Population Density, Above median days to Restaurant Closure | 395.51 ( 248.16) | 300.58 | 53.00 ( 5.323) |
|  |  |  | P=0.027 |
| Low Population Density, Median or less days to School Closure | 201.84 ( 65.06) | 223.71 | 60.22 ( 3.77) |
| Low Population Density, Above median days to School Closure | 332.30 ( 215.79) | 254.75 | 60.00 (2.00) |
| High Population Density, Median or less days to School Closure | 1146.40 (769.11) | 1061.52 | 56.00 (2.83) |
| High Population Density, Above median days to School Closure | 471.21 (404.141) | 300.58 | 53.89 ( 6.35) |
|  |  |  | P=0.047 |
| Low Population Density, Median or less days to Restrictions on gatherings | 242.67 ( 136.36) | 223.99 | 60.56 (3.57) |
| Low Population Density, Above median days to Restrictions on gatherings | 209.79 (83.96) | 223.71 | 59.00 (2.65) |
| High Population Density, Median or less days to Restrictions on gatherings | 529.95 (513.56) | 300.58 | 54.67 (4.87) |
| High Population Density, Above median days to Restrictions on gatherings | 1014.24 (726.69) | 675.19 | 54.25 (7.50) |
|  |  |  | P=0.053 |

When examining days to restaurant closure, the highest mean cumulative incidence was seen in cities with high population density and median or less days to restaurant closure, where there was a mean cumulative incidence rate of 1009.66 per 100,000. The lowest mean cumulative incidence was seen in cities with low population density and median or less days to restaurant closure, with a mean cumulative incidence rate of 188.51 per 100,000. Days of preparation was significantly different between groups. In the analysis looking at days to days to school closure, the highest mean cumulative incidence was seen in cities with high population density and median or less days to school closure, with a mean cumulative incidence rate of per 1146.40 per 100,000 people, and the lowest mean cumulative incidence was seen in cities with low population density and median or less days to school closure had a mean cumulative incidence rate of 201.84 per 100,000 people. Days of preparation was significantly different between groups. Finally, cities with low population density and above median days to restrictions on gatherings had the lowest mean cumulative incidence rate of 209.79 per 100,000 people, while the highest mean cumulative incidence rate was seen in cities with high population density and above median days to restrictions on gatherings with a rate of 1014.24 per 100,000 people. Days of preparation was not significantly different between groups.

## Discussion

The goal of this study was to quantify the effects of social distancing measures implemented by city and state governments on COVID-19 infection rates. Current commentaries on whether social distancing measures have worked have largely relied on visual representations of epidemic curves looking more or less steep (slope). However, it is crucial to gather statistical support for these observations so that solid conclusions can be made to help guide public health measures.

We found that taking longer to close schools was positively associated with cumulative incidence at the state level. The effects of social distancing measures appeared different in cities with a higher population density when compared to those with a lower population density.

Some of the findings in this study were counterintuitive, including inverse associations between cumulative incidence and days to closure of non-essential business and restrictions on gatherings. However, it seems doubtful that these measures are actually protective. This finding is more likely due to reverse causality, with locations that initially had slower growth rates initially chose not to implement measures until they absolutely needed to respond to an increased epidemic growth. Analysis is also complicated by the fact that both “non-essential business” and “gatherings” were defined very differently between locations, making a true comparison difficult. Additionally, we only used the date of the first restriction to gathering of any sort, due to the complexity of creating standard variables that can encompass the numerous variations of restrictions while still gaining meaningful statistical results.

For certain social distancing measures, such as school closure, lack of consistent results showing a protective effect on early closings should be interpreted with caution, as the small sample size for number of cities and states make it difficult to achieve statistically significant results. It also cannot be ruled out that closing of schools may not have occurred soon enough in highly populated cities to have a significant effect. Additionally, since all locations ultimately closed schools, the effect of not closing schools at all is unknown.

Our results highlight that states and cities may have different social distancing factors associated with cumulative incidence, and these factors may also be driven by additional factors such as higher population density. This suggests that not only do states and cities need individual approaches to containment of an epidemic, each location must be aware of their own structure in terms of crowding and socio-economic variables. Given the exorbitantly high rates of COVID-19 in some cities, cities cannot afford to wait for state mandates to implement social distancing measures.

The study by Courtemanche et al. may be the most similar published study to our analysis, which found no evidence that school closures or bans on large social gatherings affected growth rate. They found that closing restaurant dining rooms/bars and/or entertainment centers/gyms led to significant reductions in the growth rates. There were several aspects of their study that differed from our study, including analysis at the county level, an outcome of daily growth rate, and use of a combined variable for restaurant closure with other types of business closures. Additionally, they did not adjust for epidemics beginning at different time points in different locations or socio-demographic variables (13). The model they used is not one of comparisons between groups, however it must be recognized that the U.S. does have several sub-populations that vary in these variables.

It is important to consider that measures are needed to protect children in addition to adults. Recent data suggests that children may be susceptible to rare but severe manifestations of COVID-19 infection. As of May 5, 2020, there had been 64 suspected cases of what is being termed “Pediatric Multi-System Inflammatory Syndrome Temporally Associated with COVID-19” in New York state, with similar magnitude observed in Europe (23). There have been 4 confirmed deaths as of May 11, 2020 (24). The disease appears rare and requires further investigation, but such severe manifestations of COVID-19 in children warrants careful deliberation of the opening and closing schools.

There must also be an acknowledgement that several other variables affect cumulative incidence, including percent of Black/African American individuals in a city, percent in poverty and socioeconomic variables such as percent who own a computer, and percent who have internet access at home. Additionally, several medical conditions were associated with cumulative incidence. However, other variables that logically might be associated with cumulative incidence did not show a significant effect, including percent Hispanic, percent with hypertension, percent with heart failure and use of carpooling or public transit for work.

Many COVID-19 analyses, including the present study, suggest a disproportionate burden of illness and death among racial and ethnic minority groups, particularly the Black/African American community (25). Differences in health outcomes between racial and ethnic groups are often due to poor social determinants of health (i.e., living in densely populated and often multi-generational homes, working in essential industries often without proper insurance or sick leave, and facing institutional racism, discrimination, and stigma which undermine prevention efforts).

It is well documented that Black/African Americans have a higher risk for hypertension, type 2 diabetes, obesity, and cardiovascular diseases, which are the most common comorbidities seen in the current outbreak. These were also the most common comorbidities in previous coronavirus outbreaks such as SARS and Middle East respiratory syndrome (MERS-CoV) (26, 27, 28). It was surprising that the percent of Hispanics in a location did not show a significant association with cumulative incidence in our study, as data sources show that Hispanics (of any race) have a poverty level higher than Whites (17.5% vs 10.1%) (20, 29). Theoretically, the percent of Hispanics in a location would be associated with higher cumulative incidence as well. This finding needs to be explored in more detail.

While our study had many strengths, there were weaknesses as well. The biggest issue is the use of aggregate data from a variety of external sources. Each source has its own strength and limitations. Ideally, socio-economic variables should be evaluated by using individual level data. In terms of social distancing laws, it is unclear how each state and/or city reported their data, which may lead to a great deal of heterogeneity. Similarly, the definitions for social distancing measures were vastly different between locations. We did not distinguish between these details, and they are likely important.

Another potential limitation was our use of binary variables using median values as cutoffs for analysis. There are several ways data can be analyzed, and it is possible that there is a superior approach to analyzing social distancing variables. Despite the limitations in categorizing the data, we opted for this method because of sample size limitations which could affect statistical power. We initially conducted analyses evaluating each variable as a continuous variable but opted not to report on these results due to the complexities of reporting the effects of a change in 1 unit of a variable on a 1 point change in cumulative incidence, as is obtained from using continuous predictor variables in regression. We opted to pursue an approach that could be meaningfully understood and used by city and state governments, both in numerical and visual format.

Future studies should investigate the effects of social distancing measures in all cities, which would substantially increase sample size. Certain variables, such as use of public transit vary substantially between cities. Additionally, data can also be analyzed by new cases per day, number of hospitalizations or mortality rates, as these may show different patterns than our analysis. Despite the weaknesses in our study, the strengths of our study are the distinction between the effects of social distancing between states and cities, as well as statistical adjustment for multiple confounding variables.

Currently, it is unclear whether there will be a second wave of COVID-19 epidemics throughout the country. Limited data on wearing masks and behaviors towards gatherings suggest that failure to follow these measures may be the most challenging aspect of infection control in the U.S. A cross-sectional survey of 1034 US residents showed that 30% of people reported attending gatherings with more than 50 people and 76% of people did not wear masks outside the home (30). In contrast, a Chinese research study reported that only 3.6% of people reported going to crowded places and 2.0% reported not wearing masks outside the home (31). Ultimately, the success of government mandates will be affected by human behavior.

In conclusion, our findings demonstrate the need for a tailored, evidence-based public health response to curbing the COVID-19 pandemic. Given the immense toll the pandemic has taken on human lives in nearly every location in the U.S., all locations must take basic infection control precautions as well as social distancing measures. Areas of high risk must implement localized measures that go beyond broader state measures. While the scientific community acknowledges that there is not a single data source that will guide the pandemic response, this study demonstrates that relying solely on health services data (i.e., number of hospitalizations, tests completed, deaths, etc.) has its limitations. Government officials should be conducting robust evaluations of community mitigation strategies such as physical distancing at the state and local levels. Including an evaluation of these measures to guide reopening efforts will give officials a more holistic sense of community health and wellbeing.

Efforts should be guided by a “health in all policies’ framework, which is designed to facilitate collaboration between public health practitioners and nontraditional partners (e.g. school or transportation officials) (32). The pandemic response workforce should be trained in cultural humility, health literacy, community engagement and outreach, addressing implicit bias, and ensuring all protocols and quality improvement initiatives are systematically implemented and evaluated. Finally, a failure to follow recommended infection control strategies will likely result in a similar result with a second pandemic wave regardless of a second round of government implemented social distancing measures.

## Data Availability

All data are from publicly available sources and freely available to everyone.

https://github.com/CSSEGISandData/COVID-19/tree/master/csse_covid_19_data/csse_covid_19_time_series

https://github.com/COVID19StatePolicy/SocialDistancing

https://www.cms.gov/

https://data.census.gov/cedsci/

## Disclosures and Permissions

We confirm that this work is original. None of the authors have any conflicts of interest to disclose. This research was not funded. All data are from publicly available sources and freely available to everyone. Finally, this manuscript has been read and approved by all the authors.

## Acknowledgments

The authors recognize Daniela Fratielli, MPH Candidate at Hofstra University, for her assistance with background research.

